# “Maternal Morbidity and Medically Assisted Reproduction Treatment Types: Evidence from the Utah Population Database”

**DOI:** 10.1101/2024.08.12.24311859

**Authors:** Alina Pelikh, Ken R. Smith, Mikko Myrskylä, Michelle P Debbink, Alice Goisis

**Author notes:** Alina Pelikh is the corresponding author.

## Abstract

**Study question:** How are Medically Assisted Reproduction (MAR) treatments (Fertility enhancing drugs (FED), artificial/intrauterine insemination (AI/IUI)), assisted reproductive technology (ART) with autologous/donor oocytes) associated with maternal morbidity (MM)?

**Summary answer:** More invasive MAR treatments (ART and AI/IUI) are associated with higher risk of MM, whilst less invasive treatments are not; this relationship is partially explained by higher prevalence of multifetal gestation and obstetric comorbidities in women undergoing more invasive treatment, but the persistent association suggests subfertility itself may contribute to maternal morbidity risk.

**What is known already:** Women conceiving through MAR are at higher risk of MM, however, reported risks vary depending on the measurement of MM and data available on confounding.

**Study design, size, duration:** Birth certificates were used to study maternal morbidity among all women giving birth in Utah, U.S., between 2009 and 2017 (N=460,976 deliveries); 19,448 conceived through MAR (4.2%). The MM outcome measure included the presence of any of the following: blood transfusion; unplanned operating room procedure; admission to ICU; eclampsia; unplanned hysterectomy; ruptured uterus.

**Participants/materials, setting, methods:** Logistic regressions were estimated for the binary outcome (presence of any of the MM conditions). We assessed MM among women conceiving through MAR (overall and by type of treatment) compared to those conceiving spontaneously in the overall sample before and after adjustment for maternal socio-demographic characteristics (maternal age, family structure, level of education, Hispanic origin, parity), pre-existing maternal comorbidities (i.e., chronic hypertension, heart disease, asthma), multifetal gestation, and obstetric comorbidities (i.e., placenta previa, placental abruption, preterm delivery, cesarean delivery).

**Main results and the role of chance:** Women conceiving through MAR had higher risk of MM; however, the magnitude of the association differed depending on the type of treatment. In the unadjusted models, more invasive treatments were associated with higher odds of MM: OR 5.71 (95% CI 3.50–9.31) among women conceiving through ART with donor oocytes, OR 3.20 (95% CI 2.69–3.81) among women conceiving through ART with autologous oocytes, and OR 1.85 (95% CI 1.39–2.46) among women conceiving through AI/IUI, whereas women conceiving through FED had similar risks of MM to compared to women conceiving spontaneously (SC), OR 1.09 (95% CI 0.91–1.30). The associations between MAR and MM were largely attenuated once multifetal gestation was accounted for. After controlling for obstetric comorbidities, the associations were further attenuated, yet the coefficients remained higher among women conceiving through ART with either donor oocytes OR 1.70 (95% CI 0.95–3.04) or autologous oocytes OR 1.46 (95% CI 1.20–1.78) compared to women conceiving spontaneously. In analyses limited to singleton pregnancies, the differences in MM between women conceiving through MAR and SC were smaller in the unadjusted models. Nevertheless, women conceiving through more invasive treatments exhibited higher risk of MM. After adjusting for obstetric comorbidities, the coefficients were further attenuated and statistically insignificant for all types of treatments.

**Limitations, reasons for caution:** The data do not allow us to separate the confounding effects of subfertility on maternal morbidity from those of MAR treatments per se as there is no information on the history of previous infertility treatments or length of trying to become pregnant prior to conception. Our data also do not permit us to distinguish among different ART treatment approaches that could change certain risks (e.g. fresh or frozen embryo transfer, intracytoplasmic sperm injection, or preimplantation genetic screening via blastocyst sampling).

**Wider implications of the findings:** Our findings showing that more invasive MAR treatments are associated with higher MM suggest that subfertility could be an important unobserved factor in MM risk as it could be associated with both higher risk of MM and with undergoing more invasive procedures. Though the odds of MM were generally lower or non-significant after accounting for multifetal gestation, there remain important clinical implications because a high proportion of individuals undergoing MAR in Utah have multiple births. Therefore, the association between MAR, multifetal gestation, and MM may play a role in counselling and patient and clinician choice of MAR therapies.

**Study funding/competing interest(s):** This work was supported by European Research Council agreement n. 803958 (to A.G.). Authors have no conflict of interest to declare. MM was supported by the Strategic Research Council (SRC), FLUX consortium, decision numbers 345130 and 345131; by the National Institute on Aging (R01AG075208); by grants to the Max Planck – University of Helsinki Center from the Max Planck Society (Decision number 5714240218), Jane and Aatos Erkko Foundation, Faculty of Social Sciences at the University of Helsinki, and Cities of Helsinki, Vantaa and Espoo; and the European Union (ERC Synergy, BIOSFER, 101071773). Views and opinions expressed are, however, those of the author only and do not necessarily reflect those of the European Union or the European Research Council. Neither the European Union nor the granting authority can be held responsible for them. We thank the Pedigree and Population Resource of Huntsman Cancer Institute, University of Utah (funded in part by the Huntsman Cancer Foundation) for its role in the ongoing collection, maintenance and support of the Utah Population Database (UPDB). We also acknowledge partial support for the UPDB through grant P30 CA2014 from the National Cancer Institute, University of Utah and from the University of Utah’s program in Personalized Health and Utah Clinical and Translational Science Institute. MPD receives salary support from the March of Dimes and the American Board of Obstetrics and Gynecology as part of the Reproductive Scientist Development Program, as well as NICHD 1U54HD113169 and NIMHD 1R21MD019175-01A1.

**Trial registration number:** not applicable

## Introduction

With the increasing number of people undergoing Medically Assisted Reproduction (MAR) to treat infertility worldwide, understanding the health of children and women who conceived through MAR remains crucial. The existing literature has largely focused on the well-being of children born after MAR and established that while children conceived through MAR are more likely to have adverse perinatal outcomes (such as low birth weight or prematurity), it is likely that these are caused by maternal subfertility and high proportion of multiple births, rather than by the treatments themselves (McDonald et al. 2005; Sutcliffe and Ludwig 2007; Pandey et al. 2012; Pinborg et al. 2013; Luke et al., 2017; Berntsen et al., 2019; Goisis et al., 2019; Pelikh et al., 2022). Less attention has been given to examining the association between fertility treatments and maternal morbidity (MM), with studies showing mixed findings. Some studies show an overall increased risk of MM among women conceiving through MAR, while others report elevated risk only among specific high-risk subgroups such as women with multiple births or women with pre-existing health conditions (Belanoff et al., 2016, Wang et al., 2016; Martin et al., 2016; Dayan et al., 2019; Luke et al., 2019; Nagata et al., 2019; Korb et al., 2020; Sabr et al., 2022). As the drivers underlying these associations are complex, interconnected, and tied to multiple factors including maternal subfertility and pre-existing comorbidities, socio-demographic characteristics and obstetric complications (Lazariu et al., 2017; Kilpatrick et al., 2016; Hirshberg and Srinivas, 2017; Leonard et al., 2019; Grobman et al., 2014), it remains unclear whether MAR treatment itself poses risks for MM.

Moreover, the use of specific MAR procedures often depends on the duration of infertility and underlying infertility diagnoses along with treatment availability and cost (Henne and Bundorf, 2008; Bitler and Schmidt, 2012; Hamilton and McManus, 2012). Specifically, there is often a stepwise progression from less invasive to more invasive measures when less invasive measures do not result in ongoing pregnancy after some period of time. Due to the limited availability of large-scale data providing details on both MAR treatments and MM, only a few studies have been able to investigate whether MM differs by types of MAR treatment (Wang et al., 2016; Dayan et al., 2019; Nagata et al., 2019; Korb et al., 2020). These studies report higher MM risks among women conceiving through more invasive treatments (i.e., IVF or ICSI) compared to women who conceive spontaneously. However, the magnitude of the effects differs substantively depending on the type of the MM outcome and data employed, and findings on the effects of less invasive treatments (i.e., ovulation induction or intrauterine insemination (IUI) are mixed. This literature calls for more evidence to better understand the underlying drivers behind the association between MM and MAR. Such knowledge is crucial both for the provision and choice of treatments recommended by practitioners alongside the efforts to mitigate maternal health risks in the context of persistently high MM prevalence in the US and increasing proportion of women undergoing MAR (Tierney and Guzzo, 2023).

In this paper, we compared MM among women conceiving spontaneously and through MAR in Utah using birth certificates during the period 2009-2017. Utah has one of the highest proportion of children born through MAR of all US states - around 5% (Stanford et al., 2018, Pelikh et al., 2022) – which is comparable to some of the Nordic countries (i.e., Finland and Sweden, Wyns et al., 2021). Because birth certificate records cover all births in Utah and therefore contain a high number of women conceiving through MAR, we can investigate MM with more accuracy compared to small samples from survey data or from an individual clinic.

This study makes two key contributions. First, we analysed MM among women conceiving spontaneously and through MAR with a specific emphasis on exploring whether this association varied according to the type of treatment distinguishing between fertility enhancing drugs (FED), artificial or intrauterine Insemination (AI/IUI), and Assisted Reproductive Technology (ART) with autologous or donor oocytes. Second, we compared risks of MM among women conceiving spontaneously and through MAR before and after adjustment for a wide range of pregnancy and maternal characteristics which might confound the association between MM and MAR, such as maternal pre-existing comorbidities, socio-demographic characteristics, and for multifetal gestation and obstetric comorbidities, which might act as mediators.

## Materials and methods

### Data

This study used population-based data from the Utah Population Database (UPDB; Smith et al., 2022), which contains information from all Utah birth certificates. This study was approved by the Institutional Review Boards of the University of Utah and by the Utah Resource for Genetic and Epidemiologic Research, an administrative board overseeing access to the UPDB. The STROBE (Strengthening the Reporting of Observational Studies in Epidemiology) guidelines for cross-sectional studies were followed. Since 2009, Utah birth certificates have documented details about infertility treatments used to conceive. Through this information, we identified children conceived via specific MAR treatments – FED, AI or IUI, and ART with own and donor eggs (including donor embryos). We considered women reporting other treatments such as progesterone, metformin, and surgery for endometriosis as spontaneous conception (n=1,982), unless they also disclosed using one of the MAR procedures (n=5,134). At the time this study started, UPDB had received the birth certificate data up to 2017, marking the end of our study period.

### Study Population

The birth certificate data contain records for 469,919 deliveries registered in Utah. We excluded deliveries with missing birth order (n=247) and children born to gestational carriers (n=242). We also excluded quadruplets and quintuplets births (n=37). Further information on exclusions and missing data can be found in Figure 1. For twins and triplets, we considered one observation per delivery and controlled for multifetal gestation status. The final sample comprised 460,976 deliveries, of which 19,448 (4.2%) were conceived through MAR treatments.

**Figure 1.**
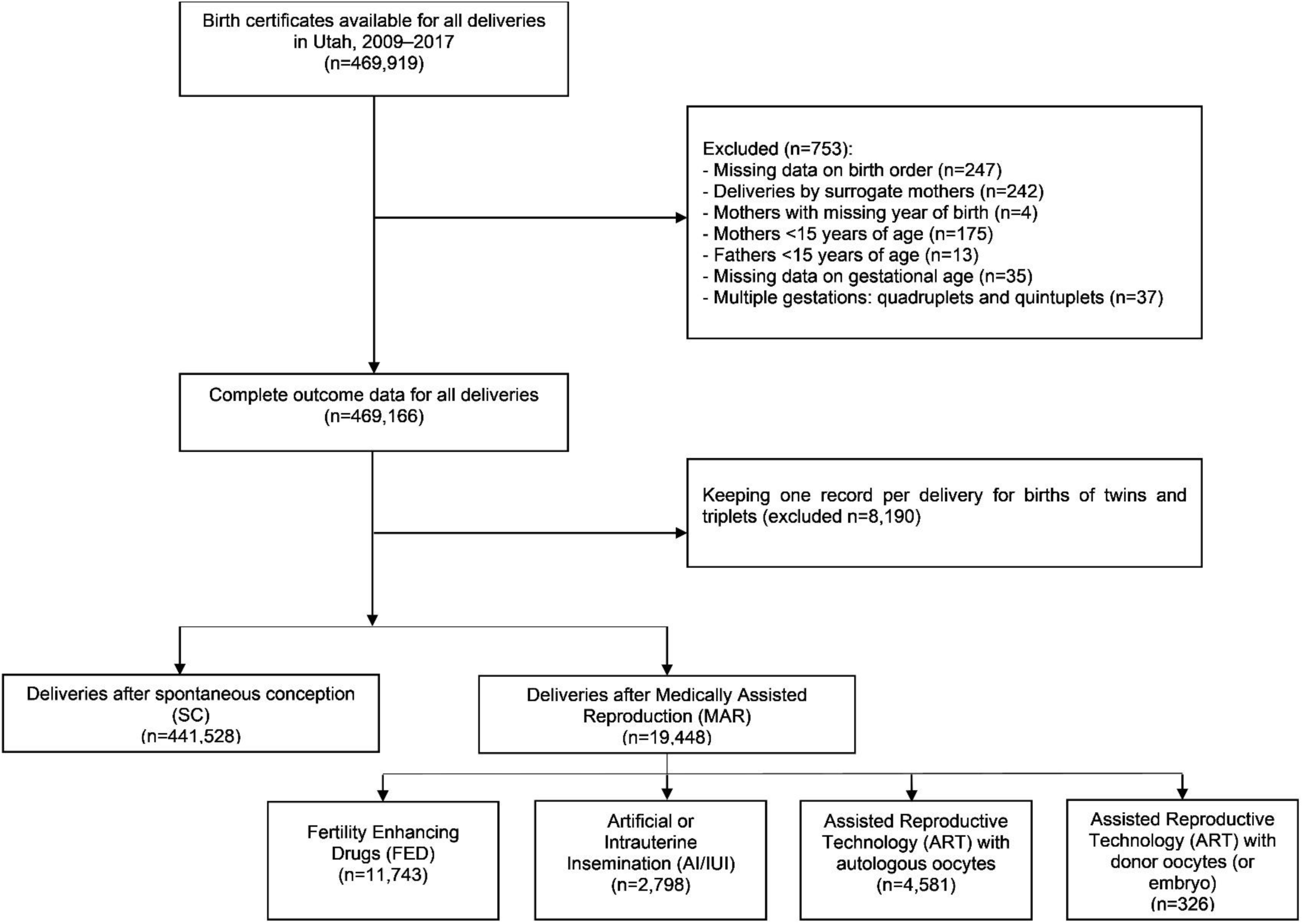
Study sample flow diagram.

### Maternal morbidity

To define maternal morbidity (MM) we used all available information registered on the birth certificate under maternal morbidities: blood transfusion; unplanned operating room procedure; admission to ICU; eclampsia; unplanned hysterectomy; ruptured uterus. Though included under the maternal morbidities heading on the birth record, we did not include 3^rd^ or 4^th^ degree perineal lacerations in our definition of MM, as these are not included in other currently accepted and validated definitions (Main et al., 2016; Snowden et al., 2021). MM was coded as a binary variable, present if any of the above events occurred. Table 1 summarises the rates of MM by mode of conception and type of MAR treatment. Given the concerns on the accuracy of blood transfusion reporting and thresholds for consideration of severity of transfusion (number of units) (Geller et al, 2004; Main et al., 2016), we performed additional sensitivity analyses excluding transfusion from the MM composite.

**Table 1.**
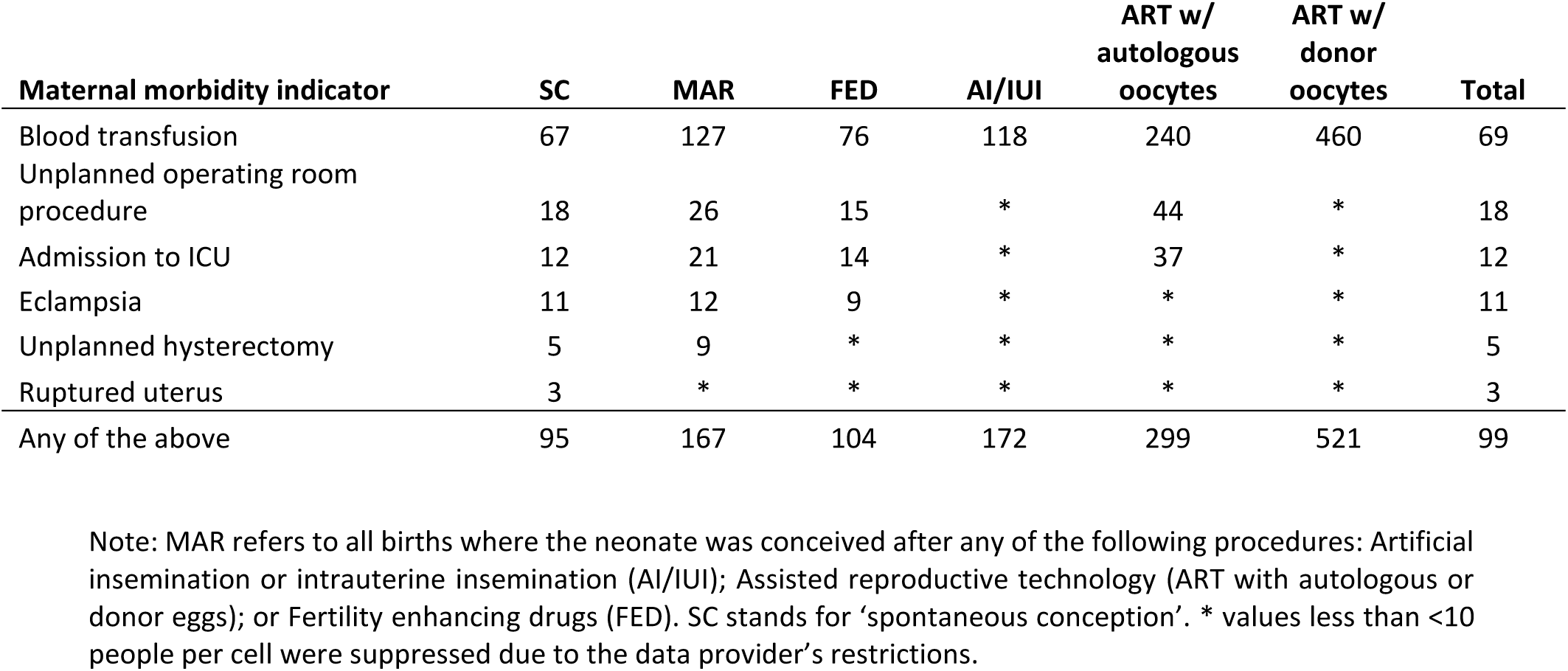
Rates of maternal morbidity per 10,000 births among women giving birth in Utah, 2009– 2017, by mode of conception and type of medically assisted reproduction treatment.

| **Maternal morbidity indicator** | **SC** | **MAR** | **FED** | **AI/IUI** | **ART w/**<br>**autologous**<br>**oocytes** | **ART w/**<br>**donor**<br>**oocytes** | **Total** |
| --- | --- | --- | --- | --- | --- | --- | --- |
| Blood transfusion | 67 | 127 | 76 | 118 | 240 | 460 | 69 |
| Unplanned operating room procedure | 18 | 26 | 15 | * | 44 | * | 18 |
| Admission to ICU | 12 | 21 | 14 | * | 37 | * | 12 |
| Eclampsia | 11 | 12 | 9 | * | * | * | 11 |
| Unplanned hysterectomy | 5 | 9 | * | * | * | * | 5 |
| Ruptured uterus | 3 | * | * | * | * | * | 3 |
| Any of the above | 95 | 167 | 104 | 172 | 299 | 521 | 99 |

Note: MAR refers to all births where the neonate was conceived after any of the following procedures: Artificial insemination or intrauterine insemination (AI/IUI); Assisted reproductive technology (ART with autologous or donor eggs); or Fertility enhancing drugs (FED). SC stands for ‘spontaneous conception’. \* values less than <10 people per cell were suppressed due to the data provider’s restrictions.

### Control variables

We considered three sets of control variables (all coded as categorical variables). The first set of factors consisted of maternal socio-demographic characteristics: maternal age (15-24; 25-29; 30-34; 35-39; 40+), marital status, the mother’s level of education (below university degree; university degree and above) and parity (first or higher-order birth). We did not include maternal race due to the very low proportion of Black, Asian, Pacific Islander and Native American women who conceived through medically assisted reproduction in Utah (i.e. <10 women per some race groups by treatment type), but we did include maternal Hispanic origin. Collectively, these characteristics could confound the association between MM and MAR (Creanga et al., 2014; Shen et al., 2005; Cabacungan et al., 2011; Berger et al., 2023).

The next set of factors was related to maternal health conditions and pre-existing comorbidities which could be associated with MM and with experiencing subfertility (Lazariu et al., 2017; Kilpatrick et al., 2016; Hirshberg and Srinivas, 2017; Leonard et al., 2019; Grobman et al., 2014). We incorporated data on asthma severity (severe and mild), chronic renal disease, chronic hypertension, heart disease severity (severe and mild), insulin-dependent diabetes, and major mental health disorder (anxiety, depression, bipolar). We could not use data on substance use, schizophrenia, rheumatic disease (rheumatoid arthritis, lupus, Sjogren’s syndrome), and non-insulin dependent diabetes available on the birth certificate due to the very low prevalence of these conditions among women conceiving via MAR. We included mother’s pre-pregnancy BMI (underweight (<18.5), healthy weight (18.5-24.9), overweight (25.0-29.9), obese (>=30)) given the demonstrated impact on both pregnancy complications and subsequent maternal health, particularly among women undergoing MAR (Dayan et al., 2015; 2018). We also accounted for maternal smoking prior to pregnancy as it is a risk factor for adverse maternal health and pregnancy outcomes (Walsh, 1994, Pollack et al., 2000). Additionally, we accounted for whether women had a history of prior caesarean deliveries, as it can influence subsequent mode of delivery and risk of MM (Clark et al., 2008; Leonard et al., 2019; Chaillet et al., 2024). We also accounted for multifetal gestation which is a common risk factor for MM (Wen et al., 2004; Luke and Brown, 2007; Gray et al., 2012; Witteveen et al., 2016).

The last group of factors was linked to obstetric comorbidities which could be related to some underlying health conditions (including subfertility) and appear to be more common among MAR conceptions but could also develop in any pregnancy posing increased risks for MM (Lazariu et al., 2017; Kilpatrick et al., 2016; Grobman et al., 2014; Hirshberg and Srinivas, 2017). Birth certificates contain information on the following conditions: placenta previa, placental abruption, preterm delivery, HELLP syndrome, pregnancy-induced hypertension, gestational diabetes, pyelonephritis, clinical chorioamnionitis, and delivery mode (cesarean delivery). Data on pyelonephritis was not included due to the very low prevalence among women conceiving via MAR. Information on haemorrhage was not included in the analysis as this condition is closely linked to blood transfusion (Lazariu et al., 2017; Kilpatrick et al., 2016; Grobman et al., 2014).

### Statistical analysis

We estimated four multivariate logistic regression models for MM. Model 1 (the baseline model) presents the unadjusted association between MAR and MM. Model 2 introduces controls for maternal socio-demographic characteristics, birth order, and pre-existing maternal comorbidities. Model 3 further includes multifetal gestation. Model 4 adds controls for obstetric comorbidities. Each model specification was estimated for all women conceiving though MAR and then differentiating by type of MAR treatment. We used clustered standard errors to account for multiple observations per woman (63.3% had one child only in the period 2009-2017).

We first compared the prevalence of MM in women conceiving via MAR to women who conceived spontaneously in the overall sample. We then restricted the analysis to singletons only to examine the associations between MM and MAR whilst removing the effects of multifetal gestation. Additionally, we estimated models which included an interaction between mode of conception and maternal pre-existing health conditions to explore whether they moderate the association between MAR and MM.

## Results

Women who conceived with FED were the largest group among all women who used MAR to conceive (n=11,743; 60.4%, Figure 1), followed by women who conceived through ART using autologous oocytes – 23.5% (n=4,581), AI or IUI – 14.4% (n=2,798), and women who conceived through ART using donor oocytes – 1.7% (n=326). Table 1 shows rates of all MM per 10,000 births, by mode of conception and type of treatments (absolute numbers are presented in Supplementary Table S1). Conditions are not mutually exclusive, therefore the total number of births with at least one condition is smaller than the sum of individual conditions. Blood transfusion was the most common MM condition reported on birth certificates (69 per 10,000 births), followed by unplanned operating room procedure and admission to ICU (18 and 12 per 10,000, respectively). Rates of the MM composite were higher among women who conceived through MAR compared to women who conceived spontaneously (167 vs 95 per 10,000 MAR or SC births, respectively). Among women who conceived through MAR, MM differed according to the level of treatment invasiveness with rates being the highest among women who conceived through ART (almost twice as high if donor oocytes were used – 521 vs 299 per 10,000 deliveries) and the lowest among women who conceived through FED (104 per 10,000 births).

The socio-demographic and health characteristics of women who conceived through MAR differed from those who conceived spontaneously (Table 2). Women who conceived through MAR were, on average, two and a half years older at the time of birth, more likely to have a degree, more likely to be married, and less likely to be of Hispanic origin. MAR children were more likely to be first-borns and multiple births (twins and triplets). Women who conceived though MAR were also more likely to experience chronic hypertension and obesity, but less likely to be smoking prior to pregnancy. The prevalence of the rest of the pre-existing comorbidities was quite similar among births to individuals using MAR or conceiving spontaneously (with some exceptions, e.g., the proportion of women with asthma and major mental health disorders was higher among women conceiving through ART with donor oocytes). Women who conceived through MAR had higher prevalence of obstetric comorbidities regardless of the type of treatments compared to women who conceived spontaneously.

**Table 2.**
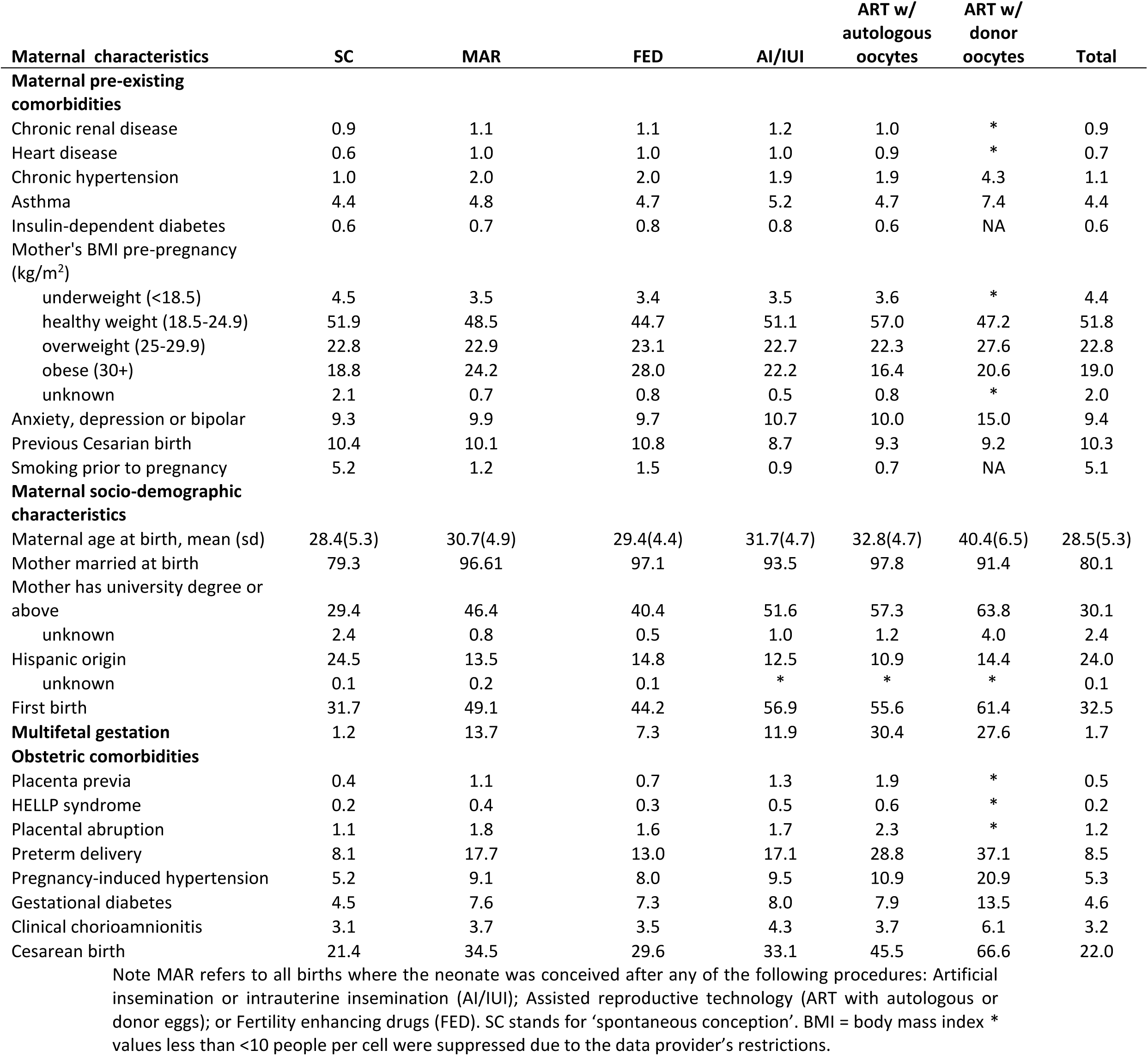
Maternal and pregnancy characteristics and obstetric comorbidities among women giving birth in Utah, 2009–2017, by mode of conception and type of medically assisted reproduction treatment (in percentages)

| Maternal characteristics | SC | MAR | FED | AI/IUI | ART w/<br>autologous<br>oocytes | ART w/<br>donor<br>oocytes | Total |
| --- | --- | --- | --- | --- | --- | --- | --- |
| **Maternal pre-existing comorbidities** |  |  |  |  |  |  |  |
| Chronic renal disease | 0.9 | 1.1 | 1.1 | 1.2 | 1.0 | * | 0.9 |
| Heart disease | 0.6 | 1.0 | 1.0 | 1.0 | 0.9 | * | 0.7 |
| Chronic hypertension | 1.0 | 2.0 | 2.0 | 1.9 | 1.9 | 4.3 | 1.1 |
| Asthma | 4.4 | 4.8 | 4.7 | 5.2 | 4.7 | 7.4 | 4.4 |
| Insulin-dependent diabetes | 0.6 | 0.7 | 0.8 | 0.8 | 0.6 | NA | 0.6 |
| Mother's BMI pre-pregnancy (kg/m<sup>2</sup>) |  |  |  |  |  |  |  |
| underweight (<18.5) | 4.5 | 3.5 | 3.4 | 3.5 | 3.6 | * | 4.4 |
| healthy weight (18.5-24.9) | 51.9 | 48.5 | 44.7 | 51.1 | 57.0 | 47.2 | 51.8 |
| overweight (25-29.9) | 22.8 | 22.9 | 23.1 | 22.7 | 22.3 | 27.6 | 22.8 |
| obese (30+) | 18.8 | 24.2 | 28.0 | 22.2 | 16.4 | 20.6 | 19.0 |
| unknown | 2.1 | 0.7 | 0.8 | 0.5 | 0.8 | * | 2.0 |
| Anxiety, depression or bipolar | 9.3 | 9.9 | 9.7 | 10.7 | 10.0 | 15.0 | 9.4 |
| Previous Cesarean birth | 10.4 | 10.1 | 10.8 | 8.7 | 9.3 | 9.2 | 10.3 |
| Smoking prior to pregnancy | 5.2 | 1.2 | 1.5 | 0.9 | 0.7 | NA | 5.1 |
| **Maternal socio-demographic characteristics** |  |  |  |  |  |  |  |
| Maternal age at birth, mean (sd) | 28.4(5.3) | 30.7(4.9) | 29.4(4.4) | 31.7(4.7) | 32.8(4.7) | 40.4(6.5) | 28.5(5.3) |
| Mother married at birth | 79.3 | 96.61 | 97.1 | 93.5 | 97.8 | 91.4 | 80.1 |
| Mother has university degree or above | 29.4 | 46.4 | 40.4 | 51.6 | 57.3 | 63.8 | 30.1 |
| unknown | 2.4 | 0.8 | 0.5 | 1.0 | 1.2 | 4.0 | 2.4 |
| Hispanic origin | 24.5 | 13.5 | 14.8 | 12.5 | 10.9 | 14.4 | 24.0 |
| unknown | 0.1 | 0.2 | 0.1 | * | * | * | 0.1 |
| First birth | 31.7 | 49.1 | 44.2 | 56.9 | 55.6 | 61.4 | 32.5 |
| **Multifetal gestation** | 1.2 | 13.7 | 7.3 | 11.9 | 30.4 | 27.6 | 1.7 |
| **Obstetric comorbidities** |  |  |  |  |  |  |  |
| Placenta previa | 0.4 | 1.1 | 0.7 | 1.3 | 1.9 | * | 0.5 |
| HELLP syndrome | 0.2 | 0.4 | 0.3 | 0.5 | 0.6 | * | 0.2 |
| Placental abruption | 1.1 | 1.8 | 1.6 | 1.7 | 2.3 | * | 1.2 |
| Preterm delivery | 8.1 | 17.7 | 13.0 | 17.1 | 28.8 | 37.1 | 8.5 |
| Pregnancy-induced hypertension | 5.2 | 9.1 | 8.0 | 9.5 | 10.9 | 20.9 | 5.3 |
| Gestational diabetes | 4.5 | 7.6 | 7.3 | 8.0 | 7.9 | 13.5 | 4.6 |
| Clinical chorioamnionitis | 3.1 | 3.7 | 3.5 | 4.3 | 3.7 | 6.1 | 3.2 |
| Cesarean birth | 21.4 | 34.5 | 29.6 | 33.1 | 45.5 | 66.6 | 22.0 |

Note MAR refers to all births where the neonate was conceived after any of the following procedures: Artificial insemination or intrauterine insemination (AI/IUI); Assisted reproductive technology (ART with autologous or donor eggs); or Fertility enhancing drugs (FED). SC stands for 'spontaneous conception'. BMI = body mass index \* values less than <10 people per cell were suppressed due to the data provider's restrictions.

Table 3 shows MAR coefficients for maternal morbidity obtained by running an unadjusted model (Model 1), after adjusting for maternal socio-demographic characteristics and pre-existing comorbidities (Model 2), multifetal gestation (Model 3) and obstetric comorbidities (Model 4). The coefficients for the control variables included in Model 2-4 are presented in Supplementary Table S2 (all births) and Supplementary Table S3 (singleton births). In Model 1, on average, women conceiving through MAR had higher odds of MM (1.76 (95% CI 1.57–1.98), but the association varied by the type of treatment. More invasive treatments (ART and AI/IUI) were, on average, associated with worse MM outcomes, while no differences were observed between women conceiving through FED compared to women conceiving spontaneously. In the unadjusted models, women conceiving through ART faced the highest odds of maternal morbidity - OR 5.71 (95% CI 3.50–9.31)) and OR 3.20 (95% CI 3.50–9.31)) among women conceiving with donor and autologous oocytes, respectively.

**Table 3.** Odds Ratios and 95% Confidence Intervals for maternal morbidity among women giving birth in Utah, 2009–2017, medically assisted reproduction compared with spontaneous conception.

|  | Model 1 (Baseline) | Model 2 (Maternal pre-existing comorbidities + socio-demographic characteristics) | Model 3 (Model 2 + multifetal gestation) | Model 4 (Model 3 + obstetric comorbidities) |
| --- | --- | --- | --- | --- |
| **a) All births** |  |  |  |  |
| MAR (*reference - SC*) | 1.76 (1.57–1.98) | 1.63 (1.45–1.84) | 1.27 (1.12–1.43) | 1.11 (0.98–1.26) |
| Type of MAR (*reference - SC*) |  |  |  |  |
| FED | 1.09 (0.91–1.30) | 1.07 (0.89–1.28) | 0.95 (0.79–1.14) | 0.87 (0.73–1.05) |
| AI/IUI | 1.85 (1.39–2.46) | 1.65 (1.24–2.19) | 1.36 (1.02–1.81) | 1.16 (0.86–1.56) |
| ART w/ autologous oocytes | 3.20 (2.69–3.81) | 2.83 (2.36–3.39) | 1.77 (1.46–2.15) | 1.46 (1.20–1.78) |
| ART w/donor oocytes | 5.71 (3.50–9.31) | 3.85 (2.29–6.46) | 2.46 (1.44–4.22) | 1.70 (0.95–3.04) |
| **b) Singleton births** |  |  |  |  |
| MAR (*reference - SC*) | 1.28 (1.11–1.48) | 1.19 (1.03–1.38) |  | 1.03 (0.89–1.20) |
| Type of MAR (*reference - SC*) |  |  |  |  |
| FED | 0.97 (0.79–1.18) | 0.95 (0.77–1.16) |  | 0.88 (0.71–1.07) |
| AI/IUI | 1.57 (1.13–2.19) | 1.40 (1.00–1.95) |  | 1.21 (0.86–1.70) |
| ART w/ autologous oocytes | 1.87 (1.42–2.46) | 1.64 (1.24–2.18) |  | 1.25 (0.93–1.66) |
| ART w/donor oocytes | 4.71 (2.50–2.87) | 3.09 (1.61–5.94) |  | 1.58 (0.74–3.38) |

Note: MAR refers to all births where the neonate was conceived after any of the following procedures: Artificial insemination or intrauterine insemination (AI/IUI); Assisted reproductive technology (ART with autologous or donor eggs); or Fertility enhancing drugs (FED). SC stands for ‘spontaneous conception’. 95% confidence intervals are presented in brackets.

After adjustment for maternal socio-demographic characteristics and pre-existing comorbidities (Model 2) the relationship between MAR and MM remained significant – OR 1.63 (95% CI 1.45–1.84). Maternal age and parity were both significantly associated with MM after being added in Model 2 and remained so even after inclusion of multifetal gestation and obstetric complications. Further adjustment for multifetal gestation (Model 3) was associated with a greater reduction in OR, yet the MM risks among women who conceived through MAR remained higher compared to women conceiving spontaneously, except for women who conceived through FED. High rates of multifetal gestation in ART contributed significantly to the MAR-MM association, with ORs attenuated to OR 2.46 (95% CI 1.44–4.22) for donor oocytes and OR 1.77 (95% CI 1.46–2.15) for autologous oocytes. These OR translate to 0.68 and 1.29 percentage points higher MM probability compared to spontaneous conception (predicted probability 0.0090 (95% CI 0.0087–0.0093)) (Table 4). Finally, after accounting for obstetric comorbidities (Model 4), the differences in odds of MM were further attenuated yet remained higher among women conceiving through ART with both donor oocytes OR 1.70 (95% CI 0.95–3.04)) and autologous oocytes OR 1.46 (95% CI 1.20–1.78) compared to women conceiving spontaneously.

**Table 4.** Predicted probabilities (adjusted for covariates) and 95% Confidence Intervals for maternal morbidity among women giving birth in Utah, 2009–2017, by mode of conception (from Model 3 in Table 3)

| Mode of conception | Predicted probability of<br>MM (95% CI) |
| --- | --- |
| SC | 0.0090 (0.0087–0.0093) |
| MAR (all) | 0.0113 (0.0100–0.0127) |
| FED | 0.0085 (0.0070–0.0100) |
| AI/IUI | 0.0121 (0.0087–0.0156) |
| ART w/autologous oocytes | 0.0158 (0.0128–0.0188) |
| ART w/donor oocytes | 0.0218 (0.0103–0.0333) |

Note: MAR refers to all births where the neonate was conceived after any of the following procedures: Artificial insemination or intrauterine insemination (AI/IUI); Assisted reproductive technology (ART with autologous or donor eggs); or Fertility enhancing drugs (FED). SC stands for ‘spontaneous conception’. 95% confidence intervals are presented in brackets. The predicted probabilities represent the average predicted probabilities after adjustments over the distribution of the categorical covariates in the data (see full list in Table 2).

In the models including only singletons, the likelihood of MM was lower in the adjusted models with all women conceiving through MAR (1.28 (95% CI 1.11–1.48) as well as among each MAR treatment type compared to women conceiving spontaneously. Similar to the results using the full sample, the associations differed by the type of treatments, with women conceiving through more invasive treatments exhibiting higher risk of MM. The association between MAR and MM was largely attenuated after controlling for obstetric comorbidities; the odds of MM remained higher than the reference group (except for women conceiving through FED) but were not statistically significant.

To investigate the moderating role of pre-existing comorbidities in the association between MAR and MM, we conducted the analyses including an interaction term between mode of conception and pre-existing health conditions (Table 5 and Supplementary Table S4). Due to the lower prevalence of MM by type of MAR treatment in the subgroups, we could only investigate the risks in the largest two groups – women conceiving through FED or through ART with autologous oocytes. In Model 1 (unadjusted), women with pre-existing comorbidities who conceived spontaneously and through MAR showed higher MM risk compared to their counterparts without pre-existing comorbidities. However, women who conceived through MAR without pre-existing comorbidities showed higher risk of MM compared to women who conceived spontaneously who had pre-existing comorbidities. A similar pattern is observed when we compared women who conceived through ART with and without pre-existing comorbidities. Adjustment for covariates in Models 2-4 attenuated but did not fully explain the group differences – women conceiving through ART with autologous oocytes were at a higher risk of MM regardless of the pre-existing comorbidities, whereas there were no differences in risks among women conceiving through FED.

**Table 5.**
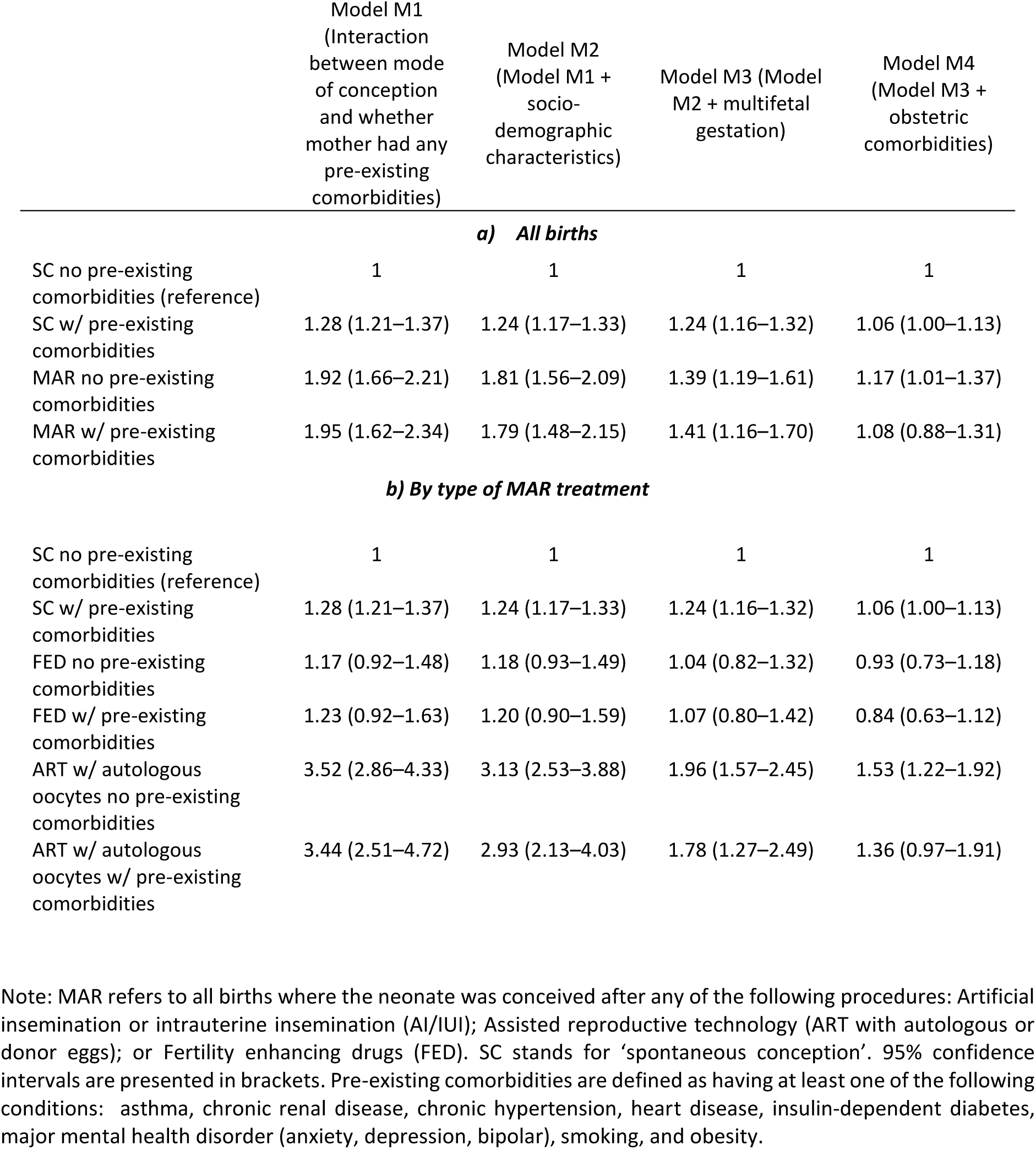
Odds Ratios and 95% Confidence Intervals for maternal morbidity among women giving birth in Utah, 2009–2017, by mode of conception and pre-existing comorbidities.

|  | Model M1<br>(Interaction<br>between mode<br>of conception<br>and whether<br>mother had any<br>pre-existing<br>comorbidities) | Model M2<br>(Model M1 +<br>socio-demographic<br>characteristics) | Model M3 (Model<br>M2 + multifetal<br>gestation) | Model M4<br>(Model M3 +<br>obstetric<br>comorbidities) |
| --- | --- | --- | --- | --- |
| **a) All births** |  |  |  |  |
| SC no pre-existing<br>comorbidities (reference) | 1 | 1 | 1 | 1 |
| SC w/ pre-existing<br>comorbidities | 1.28 (1.21–1.37) | 1.24 (1.17–1.33) | 1.24 (1.16–1.32) | 1.06 (1.00–1.13) |
| MAR no pre-existing<br>comorbidities | 1.92 (1.66–2.21) | 1.81 (1.56–2.09) | 1.39 (1.19–1.61) | 1.17 (1.01–1.37) |
| MAR w/ pre-existing<br>comorbidities | 1.95 (1.62–2.34) | 1.79 (1.48–2.15) | 1.41 (1.16–1.70) | 1.08 (0.88–1.31) |
| **b) By type of MAR treatment** |  |  |  |  |
| SC no pre-existing<br>comorbidities (reference) | 1 | 1 | 1 | 1 |
| SC w/ pre-existing<br>comorbidities | 1.28 (1.21–1.37) | 1.24 (1.17–1.33) | 1.24 (1.16–1.32) | 1.06 (1.00–1.13) |
| FED no pre-existing<br>comorbidities | 1.17 (0.92–1.48) | 1.18 (0.93–1.49) | 1.04 (0.82–1.32) | 0.93 (0.73–1.18) |
| FED w/ pre-existing<br>comorbidities | 1.23 (0.92–1.63) | 1.20 (0.90–1.59) | 1.07 (0.80–1.42) | 0.84 (0.63–1.12) |
| ART w/ autologous<br>oocytes no pre-existing<br>comorbidities | 3.52 (2.86–4.33) | 3.13 (2.53–3.88) | 1.96 (1.57–2.45) | 1.53 (1.22–1.92) |
| ART w/ autologous<br>oocytes w/ pre-existing<br>comorbidities | 3.44 (2.51–4.72) | 2.93 (2.13–4.03) | 1.78 (1.27–2.49) | 1.36 (0.97–1.91) |

Note: MAR refers to all births where the neonate was conceived after any of the following procedures: Artificial insemination or intrauterine insemination (AI/IUI); Assisted reproductive technology (ART with autologous or donor eggs); or Fertility enhancing drugs (FED). SC stands for ‘spontaneous conception’. 95% confidence intervals are presented in brackets. Pre-existing comorbidities are defined as having at least one of the following conditions: asthma, chronic renal disease, chronic hypertension, heart disease, insulin-dependent diabetes, major mental health disorder (anxiety, depression, bipolar), smoking, and obesity.

In the models with non-transfusion MM as an outcome, the risks were somewhat higher among women conceiving through ART and AI/IUI before accounting for obstetric comorbidities, but coefficients were not statistically significant. After accounting for obstetric comorbidities, the differences became both small and not statistically significant (Supplementary Table S5).

## Discussion

### Main findings

Using birth certificates from Utah, we investigated whether and how conception through different types of medically assisted reproduction treatments (fertility-enhancing drugs, AI or IUI, ART with autologous/donor oocytes) is associated with MM. In the unadjusted analyses, more invasive treatments (ART and AI/IUI) were associated with increased risks of MM, whereas women who conceived using FED had similar risks compared to women who conceived spontaneously.

The adjusted results suggest, in line with the existing literature, that the higher rates of multiple births amongst MAR conceptions play an important role in higher risk of MM amongst women who conceive via MAR. When we adjusted for multifetal gestation (in addition to maternal socio-demographic and health characteristics), the differences in MM between women conceiving through MAR and spontaneously became smaller across all MAR treatment types. Moreover, smaller differences in MM for singleton births further support the argument that the high rates of multiple birth rates associated with MAR are an important driver of these associations. Models including the sociodemographic factors also highlight the role of maternal age at birth and parity. The latter is due with the fact that most MAR children are first-borns, and nulliparity is associated with increased risk of MM. Accounting for maternal age contributed to the attenuation, particularly among women conceiving through ART with donor oocytes compared to other MAR groups due to the older profile of women in this group (mean age at birth 40.4 years vs 28.4 and 30.7 among SC and MAR-all, respectively).

Prior studies have suggested that another potential mechanism explaining the association between MAR and MM is the higher rates of pre-existing comorbidities amongst women who conceive through MAR (Lazariu et al., 2017; Kilpatrick et al., 2016; Hirshberg and Srinivas, 2017; Leonard et al., 2019; Grobman et al., 2014). The results showed that whilst pre-existing comorbidities moderated the association between MM and MAR, women who conceived via MAR remained at higher risk of MM even if they did not have pre-existing comorbidities which suggests pre-existing comorbidities only partially explain the association between MAR and MM. Another potential mechanism linking MAR and MM is the increased rate of obstetric complications such as placenta previa that occur in pregnancies conceived through MAR. After accounting for obstetric comorbidities in our models, the relationship between MAR and MM remained significant, but relatively small compared to other covariates in model 4 (Supplementary Table S2). It is difficult to conclude whether obstetric comorbidities are caused by MAR treatments, as the higher prevalence of obstetric risk factors among women conceiving through MAR could be driven by subfertility and not by the fact that they undergo MAR.

Other mechanisms which could explain the residual association between MAR and MM, such as the role of subfertility and of the MAR treatments per se, could not be directly tested. Nonetheless, our results showing that more invasive MAR treatments were associated with higher risk of MM could indirectly suggest that subfertility is an unobserved confounder as it is associated with both higher risk of MM and with undergoing more invasive MAR procedures (Belanoff et al., 2016; Luke et al., 2019; Korb et al., 2020). Although it is difficult to quantify the role played by subfertility or MAR treatments per se, adjusted results in Models 2, 3 & 4 suggest that while women undergoing MAR to conceive have higher odds of MM, the differences are small in absolute terms (Table 4) and the size of the residual MAR coefficients is smaller relative to other risk factors like maternal age, multifetal gestation, and parity (Supplementary Table S2). This could suggest that even if the MAR treatments per se play a role in MM, it is unlikely to be large.

### Strengths and limitations

This study has several strengths, including the use of high-quality vital records data to analyze MM for the entire Utah population. Such population-based data cover all births in Utah, providing a large sample of MAR conceptions and enabling more accurate MM investigation compared to smaller survey or clinic samples. Although ART usage on birth certificates may be underreported due to self-reporting (Zhang et al., 2010; Cohen et al., 2014; Luke et al., 2016), Thoma et al. (2014) found no significant ART underreporting in Utah compared to clinic-based data, unlike other US states with underreporting over 50%. found no significant underreporting in Utah compared to clinic-based data, unlike other US states with over 50% underreporting. This enhances confidence in Utah’s vital records. Additionally, we were able to distinguish between MAR treatments to examine their association with MM. MAR treatment usage in Utah’s vital records was comparable to estimates from the Pregnancy Risk Assessment Monitoring System (Stanford et al., 2018). We able accounted for a wide range of confounding factors confounding, including maternal socio-demographics, pre-existing comorbidities, pregnancy characteristics, and obstetric comorbidities. Reporting quality for maternal and child outcomes, risk factors, and obstetric history was found to be high when compared to medical charts and records across various populations (Reichman and Hade, 2001; DiGuiseppe et al, 2002; Roohan et al, 2003; Northam and Knapp, 2006; Zollinger et al, 2006; Andrade et al 2013). Overall, the high completeness and quality of Utah’s birth certificate data make it a valuable source for investigating health outcomes associated with MAR treatments.

We acknowledge some limitations of the study. Although we have utilized all available data related to MM outcomes and obstetric comorbidities from birth certificates, we could not capture the full range of indicators recommended for monitoring maternal health (i.e., pre-eclampsia, sepsis, CDC, 2015). Additionally, while we could identify four major types of MAR procedures, we could not distinguish between specific types and protocols of ART treatments which might affect MM (Nagata et al., 2019; Smith et al., 2023; Cameron et al., 2023). We also did not have information on full histories of previous fertility treatments or the precise duration or cause of infertility which prevented us from investigating in detail the effects of subfertility on maternal morbidity.

### Contributions

This study contributes to the understanding on the association between MAR and MM in multiple ways. First, using large population data we could distinguish between types of MAR treatments and show that there is a dose-response pattern as the risk of MM increases as the MAR treatments become more invasive. Women conceiving through FED face similar risks of MM to women conceiving spontaneously. Our findings are in line with previous studies reporting elevated MM among women conceiving through ART in different contexts (Belanoff et al., 2016; Martin et al., 2016; Wang et al., 2016; Dayan et al., 2019; Nagata et al., 2019; Luke et al., 2019; Korb et al., 2020; Sabr et al., 2022). However, we present novel findings by distinguishing between FED and AI/IUI as previous studies considered all non-ART infertility treatments together and while some found elevated MM risks in this group (e.g., Wang et al., 2016), others reported no difference compared to women conceiving spontaneously (Dyan et al., 2019; Nagata et al., 2019; Korb et al., 2020). Additionally, we could identify women who conceived through ART using donor oocytes, a rising group which is rarely considered in other studies due to the data availability and small sample sizes. Our findings highlight that in relation to other ART and MAR groups this group of women are at the highest risks of MM, which could be related to a range of factors including older maternal age, multifetal gestation as well as more severe form of infertility and more invasive MAR procedures. Second, we have systematically tested several potential mechanisms underlying the association between MM and MAR and showed the importance of multiple births and maternal age at birth, as well as of obstetric complications in explaining some of the association, and the limited role played by pre-existing comorbidities. This study also contributes to the discussion on the role of subfertility and MAR procedures in explaining the increased MM among women conceiving through MAR (Belanoff et al., 2016; Luke et al., 2019; Korb et al., 2020). Our findings showing that the risk of MM increases with more invasive treatments suggest that subfertility could be an important underlying factor as it is associated with both more invasive MAR procedures and MM. More evidence is needed to further investigate these associations and test these arguments.

Our main findings are relevant both for couples experiencing infertility and considering MAR treatments as well as for the public health authorities. Our findings highlight that the increased risks of MM among women conceiving through MAR are strongly associated with multifetal gestation. Multiple pregnancies could lead to increased risks for maternal health as well as adverse perinatal outcomes and health risks for children in later life which could come at high personal and public cost (Beam et al., 2020; Debbink et al., 2022). Multiple pregnancies could be caused by ovarian hyperstimulation syndrome from fertility drugs or multiple embryo transfer during ART procedures. The latter could be a preferred procedure in couples trying to maximize the chances of getting pregnant with fewer costly cycles (Swanson et al., 2020;2021). Recent evidence from the Nordic countries shows how wider state implementation of the elective single embryo transfer (eSET) policy has led to a steady decrease in the proportion of multiple pregnancies in ART over the last 15-20 years (Opdahl et al., 2020). In comparison, the national rate of eSET in the U.S. started increasing only recently – from 7% in 2009 to 67.3% in 2017 among women aged <35 year (Sunderam et al., 2012, 2020). However, the variation in practices between and within states remains high, with the rate of eSET for women aged <35 years in Utah (59.2%) lying below the national average (Sunderam et al., 2020). The lack of state-funded provision and high costs of infertility treatments could encourage the use of more invasive treatments among younger women, in particular, multiple embryo transfers, to maximize the chances to conceive. Raising awareness about health risks associated with multiple births and benefits of eSET, including similar pregnancy and live birth rate (Veleva et al., 2009; McLernon et al., 2010; Lee et al., 2016; ESHRE Guideline Group on the Number of Embryos to Transfer et al., 2024), could have a significant effect on the choice of treatments and substantially reduce the risks of adverse perinatal outcomes for women and children and associated public costs.

## Data Availability

Data regarding any of the subjects in the study has not been previously published. This study used population-based data from the Utah Population Database (UPDB; Smith et al., 2022), which contains information from all Utah birth certificates. This study was approved by the Institutional Review Boards of the University of Utah and by the Utah Resource for Genetic and Epidemiologic Research, an administrative board overseeing access to the UPDB. STROBE guidelines for a cross-sectional observational study were followed.

---

**Supplemental Table S1.** Number of maternal morbidity occurrences among women giving birth in Utah, 2009–2017, by mode of conception and type of medically assisted reproduction treatment.

| **Maternal morbidity indicator** | **SC** | **MAR** | **FED** | **AI/IUI** | **ART w/**<br>**own**<br>**eggs** | **ART w/**<br>**donor**<br>**eggs** | **Total** |
| --- | --- | --- | --- | --- | --- | --- | --- |
| Blood transfusion | 2,941 | 247 | 89 | 33 | 110 | 15 | 3,188 |
| Unplanned operating room<br>procedure | 788 | 51 | 18 | * | 20 | * | 839 |
| Admission to ICU | 513 | 41 | 17 | * | 17 | * | 554 |
| Eclampsia | 475 | 24 | 10 | * | * | * | 499 |
| Unplanned hysterectomy | 217 | 18 | * | * | * | * | 236 |
| Ruptured uterus | 112 | * | * | * | * | * | 119 |
| Any of the above | 4,216 | 324 | 122 | 48 | 137 | 17 | 4,541 |
| Total number of deliveries | 441,528 | 19,448 | 11,743 | 2,798 | 4,581 | 326 | 460,976 |

Note: MAR refers to all births where the neonate was conceived after any of the following procedures: Artificial insemination or intrauterine insemination (AI/IUI); Assisted reproductive technology (ART with autologous or donor eggs); or Fertility enhancing drugs (FED). SC stands for 'spontaneous conception'. \* values less than <10 people per cell were suppressed due to the data provider's restrictions.

**Supplemental Table S2.** Odds Ratios for maternal morbidity among women giving birth in Utah, 2009–2017, medically assisted reproduction compared with spontaneous conception (all births)

|  | MAR as one category |  |  |  | By type of MAR treatment |  |  |  |
| --- | --- | --- | --- | --- | --- | --- | --- | --- |
|  | Model 1<br>(Baseline) | Model 2<br>(Maternal pre-existing comorbidities + socio-demographic characteristics) | Model 3<br>(Model 2 + multifetal gestation) | Model 4<br>(Model 3 + obstetric comorbidities) | Model 1<br>(Baseline) | Model 2<br>(Maternal pre-existing comorbidities + socio-demographic characteristics) | Model 3<br>(Model 2 + multifetal gestation) | Model 4<br>(Model 3 + obstetric comorbidities) |
| MAR (ref: SC) | 1.76*** | 1.63*** | 1.27*** | 1.11 |  |  |  |  |
| FED |  |  |  |  | 1.09 | 1.07 | 0.95 | 0.87 |
| AI/IUI |  |  |  |  | 1.85*** | 1.65*** | 1.36* | 1.16 |
| ART w/autologous oocytes |  |  |  |  | 3.20*** | 2.83*** | 1.77*** | 1.46*** |
| ART w/donor oocytes |  |  |  |  | 5.71*** | 3.85*** | 2.46** | 1.70 |
| Chronic renal disease |  | 1.64*** | 1.65*** | 1.50*** |  | 1.64*** | 1.65*** | 1.51*** |
| Heart disease |  | 1.98*** | 1.96*** | 1.80*** |  | 1.99*** | 1.97*** | 1.81*** |
| Chronic hypertension |  | 1.25 | 1.23 | 1.13 |  | 1.25 | 1.23 | 1.13 |
| Asthma |  | 1.24*** | 1.24** | 1.18* |  | 1.24** | 1.24** | 1.18* |
| Insulin-dependent diabetes |  | 2.17*** | 2.15*** | 1.41** |  | 2.19*** | 2.17*** | 1.42** |
| Mother's BMI pre-pregnancy (ref: healthy weight (18.5-24.9) |  |  |  |  |  |  |  |  |
| underweight (<18.5) |  | 0.98 | 0.99 | 0.99 |  | 0.98 | 0.99 | 0.99 |
| overweight (25-29.9) |  | 1.00 | 1.00 | 0.95 |  | 1.01 | 1.01 | 0.95 |
| obese (30+) |  | 1.06 | 1.06 | 0.94 |  | 1.08 | 1.07 | 0.94 |
| unknown |  | 1.69*** | 1.68*** | 1.47*** |  | 1.70*** | 1.68*** | 1.47*** |
| Anxiety, depression or bipolar |  | 1.15** | 1.14** | 1.07 |  | 1.14** | 1.14** | 1.07 |
| Previous cesarian birth |  | 1.99*** | 1.99*** | 1.16** |  | 1.99*** | 1.99*** | 1.16** |
| Smoking prior to pregnancy |  | 1.08 | 1.07 | 1.01 |  | 1.08 | 1.07 | 1.01 |
| Maternal age (ref: 25-29) |  |  |  |  |  |  |  |  |
| 15-24 |  | 1 | 1.01 | 1.07 |  | 1 | 1.01 | 1.08 |
| 30-34 |  | 1.15*** | 1.13** | 1.07 |  | 1.13** | 1.13** | 1.07 |
| 35-39 |  | 1.46*** | 1.44*** | 1.27*** |  | 1.42*** | 1.41*** | 1.25*** |
| 40+ |  | 1.80*** | 1.77*** | 1.43*** |  | 1.65*** | 1.67*** | 1.36** |
| Mother married at birth |  | 0.84*** | 0.84*** | 0.87*** |  | 0.84*** | 0.84*** | 0.87*** |
| Mother has tertiary education |  | 0.94 | 0.94 | 0.98 |  | 0.93 | 0.93 | 0.98 |
| unknown |  | 1.18 | 1.17 | 1.13 |  | 1.17 | 1.17 | 1.12 |
| Hispanic origin |  | 1.01 | 1.01 | 1.01 |  | 1.01 | 1.01 | 1.02 |
| unknown |  | 0.88 | 0.86 | 0.86 |  | 0.87 | 0.85 | 0.86 |
| First birth |  | 1.60*** | 1.59*** | 1.31*** |  | 1.57*** | 1.58*** | 1.30*** |
| Multifetal gestation |  |  | 3.41*** | 1.64*** |  |  | 3.18*** | 1.56*** |
| Placenta previa |  |  |  | 5.27*** |  |  |  | 5.23*** |
| HELLP syndrome |  |  |  | 4.71*** |  |  |  | 4.71*** |
| Placental abruption |  |  |  | 3.06*** |  |  |  | 3.06*** |
| Preterm delivery |  |  |  | 2.08*** |  |  |  | 2.08*** |
| Pregnancy-induced hypertension |  |  |  | 1.37*** |  |  |  | 1.37*** |
| Gestational diabetes |  |  |  | 0.9 |  |  |  | 0.9 |
| Clinical chorioamnionitis |  |  |  | 1.65*** |  |  |  | 1.65*** |
| Cesarian birth |  |  |  | 2.30*** |  |  |  | 2.29*** |
| Constant | 0.01*** | 0.01*** | 0.01*** | 0.01*** | 0.01*** | 0.01*** | 0.01*** | 0.01*** |
| N | 460976 | 460976 | 460976 | 460976 | 460976 | 460976 | 460976 | 460976 |

Note: MAR refers to all births where the neonate was conceived after any of the following procedures: Artificial insemination or intrauterine insemination (AI/IUI); Assisted reproductive technology (ART with autologous or donor eggs); or Fertility enhancing drugs (FED). SC stands for 'spontaneous conception'. \*\*\* P < 0.001, \*\*P < 0.01, \*P < 0.05

**Supplemental Table S3.** Odds Ratios for maternal morbidity among women giving birth in Utah, 2009–2017, medically assisted reproduction compared with spontaneous conception (singleton births)

|  | MAR as one category |  |  | By type of treatment |  |  |
| --- | --- | --- | --- | --- | --- | --- |
|  | Model 1<br>(Baseline) | Model 2<br>(Maternal pre-existing<br>comorbidities + socio-demographic characteristics) | Model 4 (Model<br>3 + obstetric<br>comorbidities) | Model 1<br>(Baseline) | Model 2<br>(Maternal pre-existing<br>comorbidities + socio-demographic characteristics) | Model 4 (Model<br>3 + obstetric<br>comorbidities) |
| MAR (ref: SC) | 1.28*** | 1.19* | 1.03 |  |  |  |
| FED |  |  |  | 0.97 | 0.95 | 0.88 |
| AI/IUI |  |  |  | 1.57** | 1.40* | 1.21 |
| ART w/autologous oocytes |  |  |  | 1.87*** | 1.64*** | 1.24 |
| ART w/donor oocytes |  |  |  | 4.71*** | 3.09*** | 1.58 |
| Chronic renal disease |  | 1.62*** | 1.48** |  | 1.63*** | 1.48** |
| Heart disease |  | 2.04*** | 1.84*** |  | 2.05*** | 1.84*** |
| Chronic hypertension |  | 1.28* | 1.15 |  | 1.28* | 1.15 |
| Asthma |  | 1.25*** | 1.19* |  | 1.25*** | 1.19* |
| Insulin-dependent diabetes |  | 2.18*** | 1.39* |  | 2.19*** | 1.40* |
| Mother's BMI pre-pregnancy (ref: healthy weight (18.5-24.9)) |  |  |  |  |  |  |
| underweight (<18.5) |  | 0.98 | 0.98 |  | 0.98 | 0.98 |
| overweight (25-29.9) |  | 0.99 | 0.94 |  | 0.99 | 0.94 |
| obese (30+) |  | 1.07 | 0.93 |  | 1.07 | 0.94 |
| unknown |  | 1.73*** | 1.49*** |  | 1.73*** | 1.50*** |
| Anxiety, depression or bipolar |  | 1.16** | 1.07 |  | 1.15** | 1.07 |
| Previous cesarian birth |  | 2.01*** | 1.13* |  | 2.01*** | 1.13* |
| Smoking prior to pregnancy |  | 1.10 | 1.03 |  | 1.10 | 1.03 |
| Maternal age (ref: 25-29) |  |  |  |  |  |  |
| 15-24 |  | 0.98 | 1.05 |  | 0.98 | 1.05 |
| 30-34 |  | 1.12** | 1.05 |  | 1.11* | 1.05 |
| 35-39 |  | 1.42*** | 1.24*** |  | 1.40*** | 1.23*** |
| 40+ |  | 1.86*** | 1.49*** |  | 1.77*** | 1.45*** |
| Mother married at birth |  | 0.85*** | 0.88** |  | 0.85*** | 0.88** |
| Mother has tertiary education |  | 0.91* | 0.95 |  | 0.90** | 0.95 |
| unknown |  | 1.17 | 1.13 |  | 1.16 | 1.13 |
| Hispanic origin |  | 1.00 | 1.01 |  | 1.00 | 1.01 |
| unknown |  | 0.95 | 0.94 |  | 0.94 | 0.93 |
| First birth |  | 1.60*** | 1.31*** |  | 1.59*** | 1.30*** |
| Placenta previa |  |  | 5.35*** |  |  | 5.32*** |
| HELLP syndrome |  |  | 4.82*** |  |  | 4.80*** |
| Placental abruption |  |  | 3.05*** |  |  | 3.06*** |
| Preterm delivery |  |  | 2.11*** |  |  | 2.11*** |
| Pregnancy-induced hypertension |  |  | 1.36*** |  |  | 1.36*** |
| Gestational diabetes |  |  | 0.90 |  |  | 0.90 |
| Clinical chorioamnionitis |  |  | 1.61*** |  |  | 1.61*** |
| Cesarian birth |  |  | 2.39*** |  |  | 2.39*** |
| Constant | 0.01*** | 0.01*** | 0.01*** | 0.01*** | 0.01*** | 0.01*** |
| N | 452973 | 452973 | 452973 | 452973 | 452973 | 452973 |

Note: MAR refers to all births where the neonate was conceived after any of the following procedures: Artificial insemination or intrauterine insemination (AI/IUI); Assisted reproductive technology (ART with autologous or donor eggs); or Fertility enhancing drugs (FED). SC stands for 'spontaneous conception'. \*\*\* P < 0.001, \*\*P < 0.01, \*P < 0.05

**Supplemental Table S4.** Odds Ratios for non-transfusion maternal morbidity among women giving birth in Utah, 2009–2017, medically assisted reproduction compared with spontaneous conception (all births)

|  | MAR as one category |  |  |  | By type of treatment |  |  |  |
| --- | --- | --- | --- | --- | --- | --- | --- | --- |
|  | Model 1<br>(Baseline) | Model 2<br>(Maternal pre-existing comorbidities + socio-demographic characteristics) | Model 3<br>(Model 2 + multifetal gestation) | Model 4<br>(Model 3 + obstetric comorbidities) | Model 1<br>(Baseline) | Model 2<br>(Maternal pre-existing comorbidities + socio-demographic characteristics) | Model 3<br>(Model 2 + multifetal gestation) | Model 4<br>(Model 3 + obstetric comorbidities) |
| MAR (ref: SC) | 1.44*** | 1.29** | 1.12 | 0.96 |  |  |  |  |
| FED |  |  |  |  | 0.89 | 0.86 | 0.81 | 0.74 |
| AI/IUI |  |  |  |  | 1.92** | 1.62* | 1.46 | 1.21 |
| ART w/autologous oocytes |  |  |  |  | 2.40*** | 2.03*** | 1.55** | 1.24 |
| ART w/donor oocytes |  |  |  |  | 3.77** | 2.12 | 1.63 | 0.98 |
| Chronic renal disease |  | 2.41*** | 2.42*** | 2.19*** |  | 2.42*** | 2.42*** | 2.19*** |
| Heart disease |  | 2.77*** | 2.76*** | 2.46*** |  | 2.78*** | 2.77*** | 2.47*** |
| Chronic hypertension |  | 1.20 | 1.19 | 0.95 |  | 1.2 | 1.19 | 0.95 |
| Asthma |  | 1.33** | 1.33** | 1.27* |  | 1.33** | 1.33** | 1.27* |
| Insulin-dependent diabetes |  | 2.19*** | 2.18*** | 1.31 |  | 2.20*** | 2.19*** | 1.31 |
| Mother's BMI pre-pregnancy (ref: healthy weight (18.5-24.9)) |  |  |  |  |  |  |  |  |
| underweight (<18.5) |  | 0.89 | 0.89 | 0.86 |  | 0.89 | 0.89 | 0.87 |
| overweight (25-29.9) |  | 1.18** | 1.18** | 1.13* |  | 1.18** | 1.18** | 1.13* |
| obese (30+) |  | 1.39*** | 1.39*** | 1.26*** |  | 1.41*** | 1.40*** | 1.26*** |
| unknown |  | 2.30*** | 2.29*** | 1.94*** |  | 2.31*** | 2.30*** | 1.94*** |
| Anxiety, depression or bipolar |  | 1.08 | 1.08 | 0.99 |  | 1.08 | 1.08 | 0.99 |
| Previous cesarian birth |  | 2.09*** | 2.10*** | 1.20* |  | 2.09*** | 2.10*** | 1.20* |
| Smoking prior to pregnancy |  | 1.21 | 1.2 | 1.1 |  | 1.21 | 1.2 | 1.11 |
| Maternal age (ref: 25-29) |  |  |  |  |  |  |  |  |
| 15-24 |  | 0.91 | 0.91 | 0.98 |  | 0.91 | 0.91 | 0.98 |
| 30-34 |  | 1.33*** | 1.32*** | 1.25*** |  | 1.32*** | 1.31*** | 1.24*** |
| 35-39 |  | 1.77*** | 1.76*** | 1.53*** |  | 1.74*** | 1.73*** | 1.52*** |
| 40+ |  | 2.34*** | 2.32*** | 1.83*** |  | 2.23*** | 2.24*** | 1.80*** |
| Mother married at birth |  | 0.76*** | 0.76*** | 0.80*** |  | 0.76*** | 0.76*** | 0.80*** |
| Mother has tertiary education |  | 0.88* | 0.88* | 0.94 |  | 0.87* | 0.87* | 0.93 |
| unknown |  | 1.26 | 1.26 | 1.20 |  | 1.26 | 1.25 | 1.20 |
| Hispanic origin |  | 1.04 | 1.05 | 1.05 |  | 1.05 | 1.05 | 1.05 |
| unknown |  | 0.81 | 0.80 | 0.80 |  | 0.80 | 0.79 | 0.8 |
| First birth |  | 1.74*** | 1.74*** | 1.45*** |  | 1.72*** | 1.72*** | 1.44*** |
| Multifetal gestation |  |  | 2.23*** | 0.84 |  |  | 2.09*** | 0.8 |
| Placenta previa |  |  |  | 4.39*** |  |  |  | 4.36*** |
| HELLP syndrome |  |  |  | 2.85*** |  |  |  | 2.86*** |
| Placental abruption |  |  |  | 1.85*** |  |  |  | 1.85*** |
| Preterm delivery |  |  |  | 3.39*** |  |  |  | 3.39*** |
| Pregnancy-induced hypertension |  |  |  | 1.02 |  |  |  | 1.02 |
| Gestational diabetes |  |  |  | 0.87 |  |  |  | 0.87 |
| Clinical chorioamnionitis |  |  |  | 1.52*** |  |  |  | 1.52*** |
| Cesarian birth |  |  |  | 2.32*** |  |  |  | 2.31*** |
| Constant | 0.004*** | 0.003*** | 0.003*** | 0.002*** | 0.004*** | 0.003*** | 0.003*** | 0.002*** |
| N | 460976 | 460976 | 460976 | 460976 | 460976 | 460976 | 460976 | 460976 |

Note: MAR refers to all births where the neonate was conceived after any of the following procedures: Artificial insemination or intrauterine insemination (AI/IUI); Assisted reproductive technology (ART with autologous or donor eggs); or Fertility enhancing drugs (FED). SC stands for ‘spontaneous conception’. \*\*\* P < 0.001, \*\*P < 0.01, \*P < 0.05

**Supplemental Table S5.** Odds Ratios for non-transfusion maternal morbidity among women giving birth in Utah, 2009–2017, medically assisted reproduction compared with spontaneous conception (all births)

|  | MAR as one category |  |  |  | By type of treatment |  |  |  |
| --- | --- | --- | --- | --- | --- | --- | --- | --- |
|  | Model 1<br>(Baseline) | Model 2<br>(Maternal pre-existing<br>comorbidities<br>+ socio-demographic<br>characteristics) | Model 3<br>(Model 2 +<br>multifetal<br>gestation) | Model 4<br>(Model 3 +<br>obstetric<br>comorbidities) | Model 1<br>(Baseline) | Model 2<br>(Maternal pre-existing<br>comorbidities +<br>socio-demographic<br>characteristics) | Model 3<br>(Model 2 +<br>multifetal<br>gestation) | Model 4<br>(Model 3 +<br>obstetric<br>comorbidities) |
| MAR (ref: SC) | 1.44*** | 1.29** | 1.12 | 0.96 |  |  |  |  |
| FED |  |  |  |  | 0.89 | 0.86 | 0.81 | 0.74 |
| AI/IUI |  |  |  |  | 1.92** | 1.62* | 1.46 | 1.21 |
| ART w/autologous oocytes |  |  |  |  | 2.40*** | 2.03*** | 1.55** | 1.24 |
| ART w/donor oocytes |  |  |  |  | 3.77** | 2.12 | 1.63 | 0.98 |
| Chronic renal disease |  | 2.41*** | 2.42*** | 2.19*** |  | 2.42*** | 2.42*** | 2.19*** |
| Heart disease |  | 2.77*** | 2.76*** | 2.46*** |  | 2.78*** | 2.77*** | 2.47*** |
| Chronic hypertension |  | 1.20 | 1.19 | 0.95 |  | 1.2 | 1.19 | 0.95 |
| Asthma |  | 1.33** | 1.33** | 1.27* |  | 1.33** | 1.33** | 1.27* |
| Insulin-dependent diabetes |  | 2.19*** | 2.18*** | 1.31 |  | 2.20*** | 2.19*** | 1.31 |
| Mother's BMI pre-pregnancy (ref: healthy weight (18.5-24.9) |  |  |  |  |  |  |  |  |
| underweight (<18.5) |  | 0.89 | 0.89 | 0.86 |  | 0.89 | 0.89 | 0.87 |
| overweight (25-29.9) |  | 1.18** | 1.18** | 1.13* |  | 1.18** | 1.18** | 1.13* |
| obese (30+) |  | 1.39*** | 1.39*** | 1.26*** |  | 1.41*** | 1.40*** | 1.26*** |
| unknown |  | 2.30*** | 2.29*** | 1.94*** |  | 2.31*** | 2.30*** | 1.94*** |
| Anxiety, depression or bipolar |  | 1.08 | 1.08 | 0.99 |  | 1.08 | 1.08 | 0.99 |
| Previous cesarian birth |  | 2.09*** | 2.10*** | 1.20* |  | 2.09*** | 2.10*** | 1.20* |
| Smoking prior to pregnancy |  | 1.21 | 1.2 | 1.1 |  | 1.21 | 1.2 | 1.11 |
| Maternal age (ref: 25-29) |  |  |  |  |  |  |  |  |
| 15-24 |  | 0.91 | 0.91 | 0.98 |  | 0.91 | 0.91 | 0.98 |
| 30-34 |  | 1.33*** | 1.32*** | 1.25*** |  | 1.32*** | 1.31*** | 1.24*** |
| 35-39 |  | 1.77*** | 1.76*** | 1.53*** |  | 1.74*** | 1.73*** | 1.52*** |
| 40+ |  | 2.34*** | 2.32*** | 1.83*** |  | 2.23*** | 2.24*** | 1.80*** |
| Mother married at birth |  | 0.76*** | 0.76*** | 0.80*** |  | 0.76*** | 0.76*** | 0.80*** |
| Mother has tertiary education |  | 0.88* | 0.88* | 0.94 |  | 0.87* | 0.87* | 0.93 |
| unknown |  | 1.26 | 1.26 | 1.20 |  | 1.26 | 1.25 | 1.20 |
| Hispanic origin |  | 1.04 | 1.05 | 1.05 |  | 1.05 | 1.05 | 1.05 |
| unknown |  | 0.81 | 0.80 | 0.80 |  | 0.80 | 0.79 | 0.8 |
| First birth |  | 1.74*** | 1.74*** | 1.45*** |  | 1.72*** | 1.72*** | 1.44*** |
| Multifetal gestation |  |  | 2.23*** | 0.84 |  |  | 2.09*** | 0.8 |
| Placenta previa |  |  |  | 4.39*** |  |  |  | 4.36*** |
| HELLP syndrome |  |  |  | 2.85*** |  |  |  | 2.86*** |
| Placental abruption |  |  |  | 1.85*** |  |  |  | 1.85*** |
| Preterm delivery |  |  |  | 3.39*** |  |  |  | 3.39*** |
| Pregnancy-induced hypertension |  |  |  | 1.02 |  |  |  | 1.02 |
| Gestational diabetes |  |  |  | 0.87 |  |  |  | 0.87 |
| Clinical chorioamnionitis |  |  |  | 1.52*** |  |  |  | 1.52*** |
| Cesarian birth |  |  |  | 2.32*** |  |  |  | 2.31*** |
| Constant | 0.004*** | 0.003*** | 0.003*** | 0.002*** | 0.004*** | 0.003*** | 0.003*** | 0.002*** |
| N | 460976 | 460976 | 460976 | 460976 | 460976 | 460976 | 460976 | 460976 |

Note: MAR refers to all births where the neonate was conceived after any of the following procedures: Artificial insemination or intrauterine insemination (AI/IUI); Assisted reproductive technology (ART with autologous or donor eggs); or Fertility enhancing drugs (FED). SC stands for ‘spontaneous conception’. \*\*\* P < 0.001, \*\*P < 0.01, \*P < 0.05

## Notes

### Competing Interest Statement

The authors have declared no competing interest.

### Funding Statement

This work was supported by European Research Council agreement n. 803958 (to A.G.). MM was supported by the Strategic Research Council (SRC), FLUX consortium, decision numbers 345130 and 345131; by the National Institute on Aging (R01AG075208); by grants to the Max Planck/ University of Helsinki Center from the Max Planck Society (Decision number 5714240218), Jane and Aatos Erkko Foundation, Faculty of Social Sciences at the University of Helsinki, and Cities of Helsinki, Vantaa and Espoo; and the European Union (ERC Synergy, BIOSFER, 101071773). Views and opinions expressed are, however, those of the author only and do not necessarily reflect those of the European Union or the European Research Council. Neither the European Union nor the granting authority can be held responsible for them. We thank the Pedigree and Population Resource of Huntsman Cancer Institute, University of Utah (funded in part by the Huntsman Cancer Foundation) for its role in the ongoing collection, maintenance and support of the Utah Population Database (UPDB). We also acknowledge partial support for the UPDB through grant P30 CA2014 from the National Cancer Institute, University of Utah and from the University of Utah program in Personalized Health and Utah Clinical and Translational Science Institute. MPD receives salary support from the March of Dimes and the American Board of Obstetrics and Gynecology as part of the Reproductive Scientist Development Program, as well as NICHD 1U54HD113169 and NIMHD 1R21MD019175-01A1.

### Author Declarations

This study was approved by the Institutional Review Boards of the University of Utah and by the Utah Resource for Genetic and Epidemiologic Research, an administrative board overseeing access to the UPDB.

